# COVID-19 Critical Illness Pathophysiology Driven by Diffuse Pulmonary Thrombi and Pulmonary Endothelial Dysfunction Responsive to Thrombolysis

**DOI:** 10.1101/2020.04.17.20057125

**Authors:** Hooman D. Poor, Corey E. Ventetuolo, Thomas Tolbert, Glen Chun, Gregory Serrao, Amanda Zeidman, Neha S. Dangayach, Jeffrey Olin, Roopa Kohli-Seth, Charles A. Powell

**Author notes:** **Correspondence and request for reprints:** Hooman Poor, MD, Division of Pulmonary, Critical Care, and Sleep Medicine, Icahn School of Medicine at Mount Sinai, New York, NY, 10 E. 102^nd^ St, New York, NY 10029.

## Abstract

Critically ill COVID-19 patients have relatively well-preserved lung mechanics despite severe gas exchange abnormalities, a feature not consistent with classical ARDS but more consistent with pulmonary vascular disease. Patients with severe COVID-19 also demonstrate markedly abnormal coagulation, with elevated D-dimers and higher rates of venous thromboembolism. We present four cases of patients with severe COVID-19 pneumonia with severe respiratory failure and shock who demonstrated immediate improvements in gas exchange and/or hemodynamics with systemic tPA.

**Subject category:** 4.6 ICU Management and Outcome

## Introduction

Patients with severe COVID-19-induced respiratory failure demonstrate gas exchange abnormalities including shunt and dead-space ventilation. While patients with COVID-19 respiratory failure may fulfill the Berlin criteria for acute respiratory distress syndrome (ARDS), their syndrome is atypical in that they have relatively well-preserved lung mechanics.^1^ The marked dissociation between pulmonary mechanics and gas exchange raises the possibility of pulmonary vascular involvement.

We and others have observed a high rate of venous thromboembolism (VTE) in critically ill COVID-19 patients who otherwise lack the classic risk factors for VTE. D-dimer levels have also been noted to be elevated, and rapid rises presage cardiopulmonary decompensation. A retrospective study from China demonstrated that the use of heparin was associated with improved mortality in patients with severe COVID-19 infection and significantly elevated D-dimers.^2^ An autopsy of a patient at our institution with severe COVID-19 disease revealed numerous pulmonary microthrombi.

We present four cases of COVID-19 patients, all between 55 and 60 years old, with refractory respiratory failure requiring mechanical ventilation and shock, who demonstrated evidence of elevated dead-space ventilation. We suspected significant pulmonary micro- and/or macrothromboses as drivers of this pre-terminal state and administered systemic tissue plasminogen activator (tPA). All cases had rapid improvement in alveolar ventilation, oxygenation, and/or shock.

We obtained consent from the legally authorized representatives for all patients. This study was reviewed by the Mount Sinai Institutional Review Board and was deemed exempt.

### Case 1

Woman with a history of obesity and diabetes with COVID-19 pneumonia treated with hydroxychloroquine and ceftriaxone. She was sedated, paralyzed, and ventilated with volume-controlled ventilation (VCV) with respiratory rate (RR) 35 bpm, tidal volume (TV) 6 mL/kg IBW, FiO_2_ 60%, and PEEP 15 cmH_2_O with plateau pressure (P_pl_) of 27 cmH_2_O. Inhaled epoprostenol was administered at 25 ng/kg/min. On stable ventilator settings and epoprostenol dose, arterial blood gas (ABG) suggested significant dead-space ventilation with pH 7.12, PaCO_2_ 71 mmHg, and PaO_2_ 45 mmHg. D-dimer was elevated at 5.7 ug/mL (normal < 0.5 ug/mL). Course was complicated by vasodilatory shock requiring norepinephrine 30 mcg/min and vasopressin 2.4 units/hour, as well as acute kidney injury (AKI). Given her deterioration, she was treated with tPA 50 mg infusion over two hours. ABG at the conclusion of the infusion demonstrated marked improvement in alveolar ventilation and oxygenation with pH 7.27, PaCO_2_ 40 mmHg, PaO_2_ 78 mmHg. She was continued on a tPA infusion at 2 mg/hour with a concomitant heparin drip. Over 24 hours, her vasopressor requirement decreased to norepinephrine 4 mcg/min.

### Case 2

Woman with obesity and diabetes who required intubation in the emergency department. VCV was set at RR 30 bpm, TV 6 mL/kg IBW, FiO_2_ 70%, and PEEP 15 cmH_2_O with P_pl_ of 25 cmH_2_O. ABG suggested presence of dead-space ventilation with pH 7.33, PaCO_2_ 55 mmHg, and PaO_2_ 115 mmHg. D-dimer was elevated at 6.1 ug/mL. She had persistent shock requiring norepinephrine 15 mcg/min despite treatment with therapeutic enoxaparin. She was treated with tPA 50 mg infusion over two hours. At the conclusion of the infusion, ABG was not changed; however, she had been weaned off of vasopressors.

### Case 3

Man with obesity, hypertension, and diabetes who required initiation of mechanical ventilation in the emergency department. He was ventilated with VCV with RR 30 bpm, TV 6 mL/kg, FiO_2_ 60%, and PEEP 5 cmH_2_O with P_pl_ of 14 cmH_2_O. ABG suggested significant dead-space ventilation with pH 7.14, PaCO_2_ 107 mmHg and PaO_2_ 84 mmHg. D-dimer was elevated to 4.6 ug/mL. The clinical course was notable for AKI and refractory severe shock, requiring norepinephrine 50 mcg/min and vasopressin 2.4 units/hour. Echocardiogram demonstrated a hyperdynamic left ventricle, normal right ventricular function, and no clot in transit. Heparin drip was initiated 12 hours prior. The heparin drip was discontinued, and he was given tPA 50 mg infusion over 75 minutes. ABG at the conclusion of the infusion demonstrated improvement in alveolar ventilation, with pH 7.18, PaCO_2_ 89 mmHg, and PaO_2_ 66 mmHg, and norepinephrine dose was weaned to 7 mcg/min. Over the ensuing one hour, the patient became progressively hypoxemic and hypotensive, ultimately suffered a cardiac arrest. Echocardiogram 11 minutes into resuscitation efforts demonstrated large biventricular thrombi. The patient subsequently expired.

### Case 4

Man with obesity and hypertension. He received convalescent plasma as part of a clinical trial. Course was complicated by vasodilatory shock requiring norepinephrine 10 mcg/min and AKI. He was ventilated with VCV RR 35 bpm, TV 6 mL/kg, FiO_2_ 100%, and PEEP 16 cmH_2_O with P_pl_ of 30 cmH_2_O. ABG indicated significant dead-space ventilation with pH 7.21, PaCO_2_ 51 mmHg, PO_2_ 81 mmHg. D-dimer had increased to 6.6 ug/mL and gas exchange had not improved despite receiving a heparin drip for the preceding 24 hours. Given his deterioration, he was administered tPA 50 mg over two hours without improvement in gas exchange or hemodynamics. He was restarted on a heparin drip and a tPA drip was initiated at 2 mg/hr. After 12 hours, ABG demonstrated marked improvement in oxygenation with pH 7.27, PaCO_2_ 51 mmHg, and PaO_2_ 140 mmHg (Figure 1).

**Figure 1.**
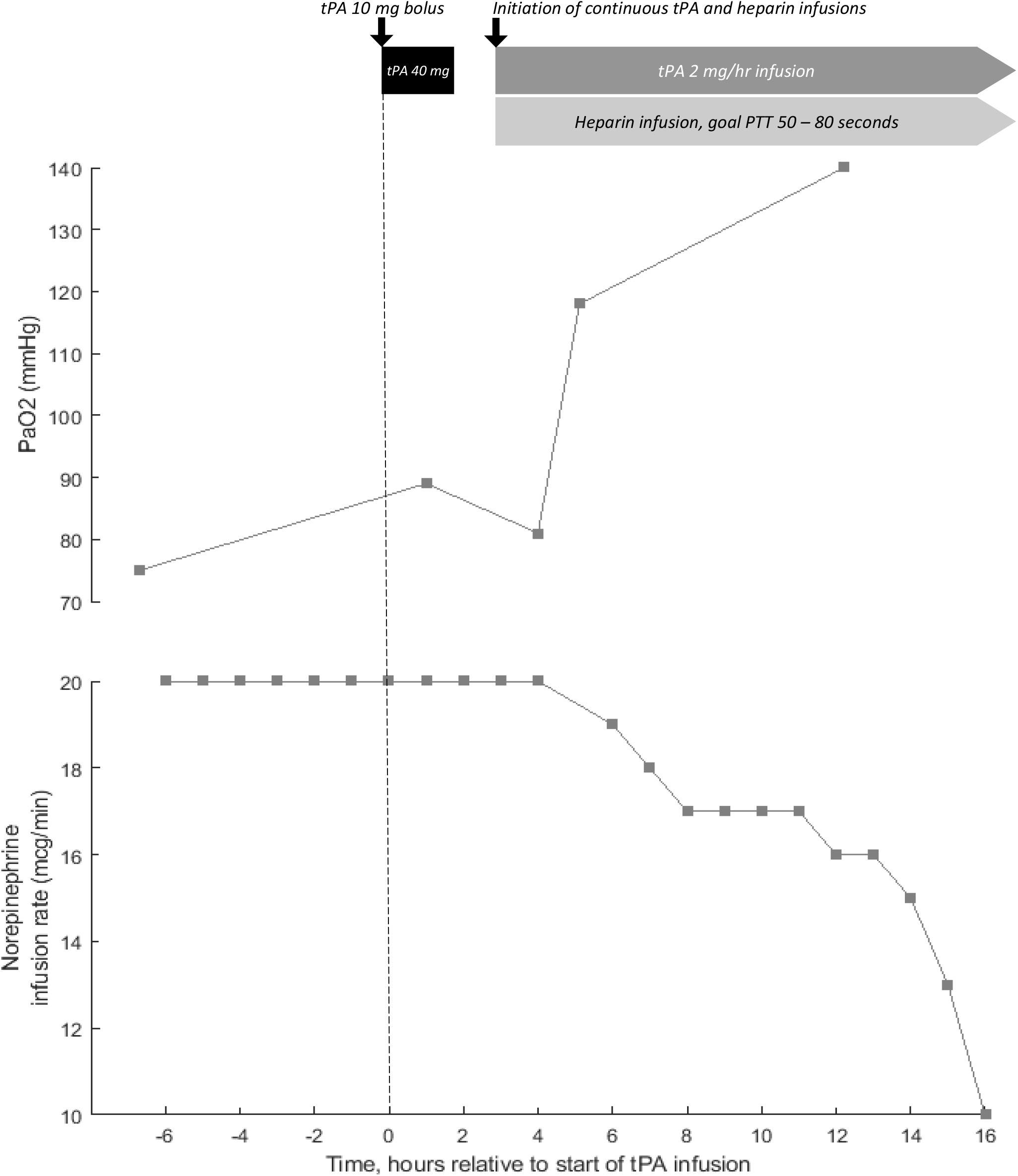
Time course of PaO_2_ and norepinephrine dose relative to two different tPA infusion strategies seen in Case #5. Administration of tPA 10 mg bolus followed by 40 mg infusion over two hours in the absence of concomitant heparin did not result in change in PaO_2_ or norepinephrine dose. Low-dose tPA with concomitant heparin demonstrated significant improvements in PaO_2_ and shock.

## Discussion

We suspect that the primary mechanism by which COVID-19 causes respiratory failure is pulmonary endothelial dysfunction with diffuse, heterogeneously distributed pulmonary microthrombi in some lung regions, and significant pulmonary vascular dilatation in other lung regions. These simultaneous abnormalities could explain the combined dead-space and shunt physiology, as well as the preserved pulmonary hemodynamics and normal right ventricular function reported previously.^3^ The coexistence of obliterative lesions and vasodilatory regions is reminiscent of the pathophysiology seen in cirrhotic patients with portopulmonary hypertension (obliterative) and hepatopulmonary syndrome (vasodilatory).^4^ The improvement in oxygenation noted with increased PEEP in COVID-19 “ARDS” may be attributable to decreased cardiac output, leading to decreased shunt fraction rather than to alveolar recruitment.^1^

Autopsy studies from the SARS outbreak of the early 2000s, caused by SARS-CoV-1 virus, have demonstrated pulmonary thrombi, pulmonary infarcts, and microthrombi in other organs.^5-8^ It appears that SARS-CoV-2 is causing similar pathophysiological derangements. Although microthrombi are present in sepsis and classic forms of ARDS, they are unlikely to be the principal cause of respiratory failure and organ dysfunction.^9-10^ In COVID-19 pneumonia, the thrombi may play a direct and significant role in gas exchange abnormalities and in multisystem organ dysfunction. The preserved lung compliance noted early in the course of COVID-19 patients with bilateral airspace opacities suggests that the observed pulmonary infiltrates could represent areas of pulmonary infarct and hemorrhage. In our series, disseminated intravascular coagulation was not the cause of microthrombi as all four patients had normal platelet levels without demonstration of hemolysis, despite elevated D-dimer levels. The high prevalence of obesity, hypertension, and diabetes in patients with severe COVID-19 pneumonia may point to an underlying susceptibility to endothelial injury and dysregulation in this metabolic syndrome.

Thrombolysis in this case series had an immediate, physiological impact that variably improved alveolar ventilation, oxygenation, and shock. Thrombolysis improves alveolar ventilation by restoring blood flow to previously occluded regions. This redistribution would reduce blood flow to vasodilated vessels, decreasing the shunt fraction and improving oxygenation. The improvement in shock may be multifactorial, but could be secondary to reperfusion of other ischemic organs that have microthrombi (e.g. the kidneys, as all five cases had AKI), leading to an improvement in the overall inflammatory and vasodilatory state.

These four cases had respiratory failure early in their COVID-19 course and had evidence of the “pulmonary vascular” phenotype (normal compliance, increased dead-space, elevated D-dimers). It may be prudent to consider full systemic anticoagulation for earlier disease to possibly prevent or mitigate progression of the syndrome. As this phenotype progresses, and patients develop severe progressive cardiopulmonary compromise, therapeutic anticoagulation alone may not be effective and thus systemic thrombolysis may be beneficial; Cases 3 and 4 had been receiving therapeutic anticoagulation prior to tPA administration without noticeable improvement. A second more classic “ARDS” phenotype may exist as a discrete entity or as part of a spectrum of disease with low compliance from continued lung injury due to mechanical ventilation or COVID-19 induced lung injury.

The pathophysiology of COVID-19 severe respiratory failure may be driven by pulmonary vascular endothelial dysfunction and thrombosis that responds to thrombolysis and anticoagulation. These therapeutic approaches should be considered in the management of COVID-19 patients and must be further examined in clinical research studies.

## Data Availability

This manuscript is a case series of five patients.

## Author’s contributions

H.P.: concept, clinical care, data gathering, interpretation, drafting of the manuscript; C.E.V.: concept, interpretation, revision of the manuscript; T.T.: clinical care, data gathering, interpretation, revision of the manuscript; G.C.: clinical care, data gathering, interpretation, revision of the manuscript; G.S.: interpretation, revision of the manuscript; A.Z.: interpretation, revision of the manuscript; N.D.: clinical care; J.O.: interpretation, revision of the manuscript; R.K.S.: clinical care, interpretation, revision of the manuscript; C.A.P.: concept, clinical care, interpretation, revision of the manuscript

## References

1. Gattinoni L, Coppola S, Cressoni M, Busana M, Chiumello D. Covid-19 Does not Lead to a “Typical” Acute Respiratory Distress Syndrome. Am J Respir Crit Care Med [online ahead of print] 30 Mar 2020; DOI: 10.1164/rccm.202003-0817LE.

2. Tang N, Bai H, Chen X, Gon J, Li D, Sun Z. Anticoagulant treatment is associated with decreased mortality in severe coronavirus disease 2019 patients with coagulopathy. J Thromb Haemost [online ahead of print] 27 Mar 2020; DOI: 10.1111/jth.14817.

3. Kelly G, Alexander P, Clifford M. Pediatrica Intensiva Podcast, 2.2 The Frontlines of COIVD19: Italian intensivists Gio Colombo & Lorenzo Grazioli 2 weeks into their enormous epidemic. 21 Mar 2020. Available from: http://pedsintensiva.libsyn.com/22-the-frontlines-of-covid19.

4. Porres-aguilar M, Altamirano JT, Torre-delgadillo A, Charlton MR, Duarte-rojo A. Portopulmonary hypertension and hepatopulmonary syndrome: a clinician-oriented overview. Eur Respir Rev. 2012;21(125):223–33.

5. Chong PY, Chui P, Ling AE, et al. Analysis of deaths during the severe acute respiratory syndrome (SARS) epidemic in Singapore: challenges in determining a SARS diagnosis. Arch Pathol Lab Med. 2004;128(2):195–204.

6. Hwang DM, Chamberlain DW, Poutanen SM, Low DE, Asa SL, Butany J. Pulmonary pathology of severe acute respiratory syndrome in Toronto. Mod Pathol. 2005;18(1):1–10.

7. Ng KH, Wu AK, Cheng VC, et al. Pulmonary artery thrombosis in a patient with severe acute respiratory syndrome. Postgrad Med J. 2005;81(956):e3.

8. Xiang-hua Y, Le-min W, Ai-bin L, et al. Severe acute respiratory syndrome and venous thromboembolism in multiple organs. Am J Respir Crit Care Med. 2010;182(3):436–7.

9. Wilde JT, Roberts KM, Greaves M, Preston FE. Association between necropsy evidence of disseminated intravascular coagulation and coagulation variables before death in patients in intensive care units. J Clin Pathol. 1988;41(2):138–42.

10. Tomashefski JF, Davies P, Boggis C, Greene R, Zapol WM, Reid LM. The pulmonary vascular lesions of the adult respiratory distress syndrome. Am J Pathol. 1983;112(1):112–26.

